# Epigenetic inflammation signatures and lung cancer risk among never-smoking women: a nested case-control study

**DOI:** 10.64898/2026.04.27.26351864

**Authors:** Mohammad L. Rahman, Aishani Gargapati, Lauren M. Hurwitz, Wei Hu, Alexander P. Keil, Charles E. Breeze, Anil Chaturvedi, Jianxin Shi, Qiuyin Cai, Gong Yang, Jirong Long, Yu-Tang Gao, David C. Christiani, Nathaniel Rothman, Wei Zheng, Xiao-Ou Shu, Jason Y.Y. Wong, Qing Lan

**Author notes:** To whom correspondence should be addressed: **Mohammad L Rahman**, MD, ScD, MPH, Occupational & Environmental Epidemiology Branch, Division of Cancer Epidemiology and Genetics, National Cancer Institute, NIH, 9609 Medical Center Drive, Rm. 6E132, Rockville, MD 20850, USA. 9. University of Texas at Tyler School of Medicine, Tyler, TX. These authors co-supervised this work (Wei Zheng, Xiao-Ou Shu, Jason Y.Y. Wong, Qing Lan).

## Abstract

**Introduction:** Chronic inflammation has been implicated in lung carcinogenesis. Prospective studies have linked higher circulating C-reactive protein (CRP), an acute-phase inflammation marker, to higher lung cancer risk in predominantly smoking populations but lower risk in never smokers. We evaluated DNA methylation–based inflammation risk scores (DNAm-IRSs), which may capture longer-term immune-inflammatory and exposure-related biology, with lung cancer risk among never smokers.

**Methods:** We evaluated six DNAm-IRSs, including four CRP-based scores (IRS_Ligthart_, IRS_Wielscher_, IRS_Linear_Hillary_, IRS_Elnet_Hillary_), in 683 risk-set-sampled case-control pairs nested in the Shanghai Women’s Health Study (n=74,941). We estimated hazard ratios (HRs) and 95% confidence intervals (CIs) using conditional logistic regression. We examined DNAm-derived leukocyte composition and circulating immune-inflammatory proteins to characterize DNAm-IRS biology.

**Results:** Circulating CRP correlated positively with IRS_Ligthart_ (r=0.19), IRS_Wielscher_ (r=0.13), and IRS_Elnet_Hillary_ (r=0.30), but inversely with IRS_Linear_Hillary_ (r=-0.02). Per standard deviation increase, IRS_Ligthart_ was associated with lower lung cancer risk (HR=0.85, 95% CI: 0.76–0.95), and IRS_Wielscher_ with lower risks of lung cancer (HR=0.87, 95% CI: 0.77–0.97) and adenocarcinoma (HR=0.83, 95% CI: 0.71–0.97). Associations persisted after adjustment for leukocyte composition and strengthened after adjustment for DNAm pack-years, an epigenetic smoking index that may capture combustion-related exposures beyond active smoking. Inverse associations were more evident among women with lower DNAm pack-years, although formal interaction tests were not statistically significant. Both scores were positively associated with acute-phase inflammation, IFN-γ/effector trafficking, and higher CD8+ T-cell proportions.

**Conclusions:** Among never smokers, selected CRP-related DNAm-IRSs were associated with lower lung cancer risk and were linked to immune features consistent with antitumor activity.

## Introduction

Lung cancer remains the leading cause of cancer mortality worldwide. A substantial proportion of cases occur in never-smokers, estimated at ∼15% in men and ∼53% in women globally,[1] yet the etiologic drivers in this population remain incompletely understood. Unlike smoking-associated lung cancer, carcinogenesis in never-smokers likely reflects heterogeneous environmental exposures and host-response processes,[2] among which chronic inflammation has emerged as a key candidate mechanism.[3]

Inflammation plays a complex and context-dependent role in carcinogenesis: it can promote tumor development through oxidative stress and DNA damage or, conversely, support antitumor immunity and antigen presentation.[4] However, the specific inflammatory profiles that influence lung cancer risk in never-smokers, and their etiologic relevance, remain poorly characterized.

C-reactive protein (CRP), a widely used circulating marker of systemic inflammation, has been prospectively associated with higher lung cancer risk in predominantly smoking populations.[5-8] However, prior subgroup analysis restricted to never smokers were substantially underpowered (n <85 cases),[5-7] whereas a large, pooled analysis from the Lung Cancer Cohort Consortium (1,305 cases) reported an inverse association.[8] Similarly, we observed an inverse association between circulating CRP measured in baseline and lung cancer risk among never-smoking women (248 cases) in the Shanghai Women’s Health Study (SWHS).[9]

Interpretation of associations between lung cancer and CRP is limited by its acute-phase kinetics and short plasma half-life (∼19 hours),[10] making single measurements susceptible to transient perturbations (e.g., infection or injury) and potentially inadequate for capturing chronic inflammatory processes underlying long-term disease risk. Such within-person variability and measurement error may attenuate associations and contribute to heterogeneity across populations differing in smoking prevalence, air pollution, adiposity, and other determinants of systemic inflammation.

DNA methylation (DNAm)–based inflammation risk scores (IRS) offer epigenetic proxies that may better capture long-term inflammatory states. Large epigenome-wide association studies have identified CpG signatures with CRP associations that replicate across populations,[11] enabling construction of DNAm-derived CRP predictors.[11-14] Rather than simple surrogates for circulating CRP, these scores function as composite markers of chronic immune-inflammatory and exposure-related processes.[11, 14] However, evidence linking these scores to lung cancer incidence remains limited: a DNAm-derived CRP score showed no association in smoker-prevalent Campaigns for Lung Cancer Prevention and Early Detection (CLUE I/II) cohorts,[13] whereas DNAm proxies for other markers, including interleukin-6 (IL-6) and methylation-derived neutrophil-to-lymphocyte ratio (mdNLR),[15, 16] have demonstrated more consistent positive associations.[9, 13, 17-19]

Inflammatory markers, either circulating proteins or DNAm-based scores, may reflect different biologic processes depending on exposure context. In individuals without major inflammatory stressors, these markers may better capture baseline inflammatory tone or host immune state, whereas in exposed populations, they may instead reflect exposure-induced inflammatory responses. This distinction is critical when the same exposures also influence lung cancer risk. Smoking is a clear example: it increases systemic inflammation, alters DNAm, and independently drives cancer formation through mutagenic and related pathways.[20-22] The opposite directions reported for circulating CRP and lung cancer associations in predominantly smoking populations versus never-smokers are consistent with this possibility. Other exposures, including combustion-related air pollution, may act similarly. In such settings, CRP-related markers may partly function partly as proxies for carcinogenic exposures rather than as purely indexing intrinsic inflammatory biology, creating potential confounding and exposure-dependent heterogeneity if those exposures are inadequately captured. Consistent with this framework, in the smoker-dominant CLUE I/II cohorts, strict adjustment for smoking using DNAm-derived smoking pack-years, an epigenetic smoking index, revealed an inverse association between a DNAm-derived CRP score and lung cancer,[13] suggesting that the etiologic relevance of inflammation may differ when disentangled from co-occurring carcinogenic exposures.

Importantly, DNAm pack-years may capture broader combustion-related exposures beyond active smoking. In never-smoking women exposed to household air pollution from indoor coal combustion, DNAm pack-years was positively correlated with cumulative exposure to a cluster of 36 polycyclic aromatic hydrocarbons (PAHs; Spearman *ρ* = 0.31; P = 5.1×10□□), including 5-methylcrysene, a potent lung carcinogen.[23] In the same population, household air pollution was associated with GrimAge acceleration, an epigenetic aging metric that incorporates DNAm pack-years.[24, 25] Likewise, traffic- and wildfire-related particulate matter exposures have been linked to GrimAge acceleration and canonical smoking-related CpG sites annotated to the aryl hydrocarbon receptor repressor (*AHRR*) and coagulation factor II thrombin receptor-like 3 (*F2RL3*) genes.[26-28] Together, these findings support the use of DNAm pack-years as a proxy for cumulative combustion-related exposure in never smokers and that it may help address residual or unmeasured confounding in analyses of inflammatory markers and lung cancer risk.

Here, we examined associations between DNAm-IRS derived from pre-diagnostic blood DNA methylation profiles and lung cancer risk among never-smoking women. This study addresses three key gaps. First, it leverages multiple DNAm-IRS as potentially more stable and integrative indicators of chronic inflammatory biology than single protein measurements. Second, beyond restricting analyses to never-smokers, it evaluates the robustness of associations after additional adjustment for DNAm pack-years, a proxy for cumulative combustion-related exposures, and assesses potential effect modification by this marker. Third, it integrates circulating immune-inflammatory markers and DNAm-derived immune cell composition to characterize the biological phenotypes indexed by DNAm-IRS.

## Material and Methods

### Study population and design

This nested case–control study was conducted within the Shanghai Women’s Health Study (enrollment 1997–2000; n=74,941), a population-based prospective cohort in Shanghai, China, previously described. [29] Participants completed baseline and follow-up in-person interviews capturing demographics, occupational and environmental exposures and lifestyle factors; cohort participation was 92.7%.

Incident cancers were ascertained through in-person follow-up surveys every 2–3 years and annual linkage with the Shanghai Cancer Registry and the Vital Statistics Unit. Eligible cases were incident lung cancers among lifetime never-smokers, defined according to the U.S. Centers for Disease Control and Prevention criterion of <100 cigarettes smoked over the lifetime. For each case, matched controls were selected using risk-set sampling (incidence density sampling) from cohort members who were alive and cancer free at the time of the case diagnosis, with matching on date of birth (±2 years) and date of blood collection (±3 months).

Lung cancer was identified using International Classification of Diseases, Ninth Revision (ICD-9) codes 162.1-162.9 and further classified by histology (adenocarcinoma, non-adenocarcinoma, or unclassified) using ICD-O-2 morphology codes (80003, 80413, 80703, 81403, 82403, 82603, 84803, 85503, 85603). Cases were diagnosed between 2000 and 2016 (median follow-up 10.1 years; range 0–17 years).

All participants provided written informed consent, and study protocols were approved by the institutional review boards of all participating institutions.

### DNA methylation profiling and derivation of DNAm-IRS

Genomic DNA was extracted from buffy coat and profiled at the NCI Cancer Genomics Research Laboratory using the Illumina Infinium MethylationEPIC BeadChip. Raw IDAT files were processed in R using Bioconductor packages and the ChAMP pipeline.[30] Standard probe-level quality control excluded probes with poor detection, low bead count, non-CpG annotation, SNP overlap, multi-mapping, or sex chromosome localization. After filtering, 692,282 CpGs remained for analysis. Missing values were imputed by k-nearest neighbors, beta values were normalized using Beta Mixture Quantile dilation, and batch effects were corrected using ComBat.[31-33] Leukocyte composition were estimated from normalized methylation data using the Houseman method.[34] The final analytical sample included 683 case-control pairs.

DNAm-based scores were calculated using published algorithms, restricted to CpGs retained after quality control and preprocessing. Specifically, we derived four CRP-based scores: IRS_Ligthart_ (52 CpGs), IRS_Wielschar_ (1,333 CpGs), IRS_Linear_Hillary_ (32,196 CpGs), IRS_Elnet_Hillary_ (1,334 CpGs), and one IL-6-based score: IRS_Stevenson_ (34 CpGs) as well as a methylation-derived neutrophil-to-lymphocyte ratio (mdNLR), using published algorithms.[11, 12, 14, 16] mdNLR was log-transformed, and all DNAm-based indices were standardized before analysis. DNAm pack-years was calculated as a weighted sum of smoking-associated CpG sites (1,580 CpGs) using published regression coefficients for pack-years as weights.[20]

### Circulating immune–inflammatory markers

A total of 61 circulating immune-inflammatory markers were measured using Luminex bead-based assays and previously examined in relation to lung cancer risk.[9] Markers meeting prespecified analytic criteria (≥90% detectable, within-batch coefficient of variation ≤15%, and intraclass correlation coefficient ≥0.80) were grouped a priori into functional modules based on cytokine or chemokine families and established immunologic roles (**Table S1, S2**). Immune module analyses were restricted to participants with available protein data (201–265 cases and 185–222 controls, depending on the marker). Protein concentrations were log-transformed and standardized to z scores, and module scores were defined as the mean of the standardized values of component proteins. For multiprotein modules, scores were calculated only when all component proteins were available; single-protein modules were represented by the standardized value of the corresponding marker.

### Statistical analysis

Weighted Spearman correlations were estimated among DNAm-IRS, circulating CRP and IL-6 levels, and DNAm pack-years using inverse probability weights based on lung cancer prevalence in the source cohort. To assess associations between DNAm-IRS and lung cancer, we first examined the shape of the associations using generalized additive models with penalized splines adjusted for age and body mass index (BMI). Because these analyses provided little evidence of nonlinearity (**Figure S1**), all DNAm-IRS were modeled continuously per 1-standard deviation (SD) increase.

Associations between standardized DNAm-IRS and lung cancer risk were estimated using conditional logistic regression with stratification by matched set to account for the nested case-control design. Under risk-set sampling, these estimates are interpretable as hazard ratios (HRs) and are reported with 95% confidence intervals (CIs). The base model adjusted for age and BMI. A second model additionally adjusted for estimated leukocyte compositions to assess whether associations were independent of immune cell composition; this adjustment was not applied to mdNLR because it is derived from leukocyte subtypes. A third model further adjusted for DNAm pack-years as a proxy for cumulative combustion-related exposure. To reduce potential reverse causation, we repeated analyses after excluding cases diagnosed within 2 years of blood collection. Analyses were conducted for overall lung cancer and adenocarcinoma.

For overall lung cancer, additional analyses were restricted to histologically confirmed cases. To evaluate heterogeneity by timing of diagnosis, analyses were further stratified by the median follow-up time from blood draw to diagnosis (10.1 years). Potential effect modification by combustion-related exposures was assessed by stratifying analyses according to high versus low DNAm pack-years, defined using the median among controls. In addition, multiplicative interaction between each DNAm-IRS and continuous DNAm pack-years was evaluated by including cross-product terms. Multiple testing across DNAm-IRS was controlled using the Benjamini-Hochberg false discovery rate procedure.

To provide biological context for the DNAm-IRS associations with lung cancer, we evaluated associations of selected DNAm-IRS with functional immune-inflammatory module scores using linear regression with adjustment sets paralleling those used in the main analyses. We also tested effect modification by DNAm pack-years. In addition, we examined associations between DNAm-IRS and estimated leukocyte compositions, with leukocyte compositions logit-transformed before analysis. Additional details are provided in the Supplementary Methods.

## Results

Participant characteristics are presented in **Table 1**. Among cases, about 80% were histology-confirmed and 57.5% were lung adenocarcinoma. The CRP-based scores were generally intercorrelated (r=0.18 to 0.85) but showed mixed correlations with circulating CRP, with positive correlations for IRS_Ligthart_ (r=0.19), IRS_Wielscher_ (r=0.13), and IRS_Elnet_Hillary_ (r=0.30) and a near-null inverse correlation for IRS_Linear_Hillary_ (r=-0.02). The IL-6-based score, IRS_Stevenson_ was weakly correlated with circulatory IL-6 (r=0.13). Several DNAm-IRSs were also positively correlated with DNAm pack-years, most notably IRS_Ligthart_ (r =0.61) and IRS_Wielscher_ (r=0.46) (**Table S3**).

**Table 1.** Participant characteristics by lung cancer case-control status.

| Characteristics | Controls (n= 683) | Cases (n= 683) |
| --- | --- | --- |
| Age at baseline, mean ± SD | 56.36 ± 8.88 | 56.24 ± 8.85 |
| Body mass index, mean ± SD | 24.64 ± 3.45 | 24.32 ± 3.45 |
| Years of diagnosis from blood collection |  | 10.11 (5.88, 13.48) |
| DNAm pack-years, $\times 10^6$ | -6.53 (-6.65, -6.41) | -6.51 (-6.65, -6.38) |
| DNAm-IRS scores |  |  |
| IRS<sub>Ligthart</sub> | -0.03 (-0.04, -0.03) | -0.03 (-0.04, -0.03) |
| IRS<sub>Wielscher</sub> | 64.44 (43.19, 91.18) | 59.99 (40.75, 82.23) |
| IRS<sub>Linear_Hillary</sub> | -3290.62 (-3922.56, -2487.09) | -3350.67 (-3916.51, -2660.96) |
| IRS<sub>Elnet_Hillary</sub> | -0.12 (-0.13, -0.12) | -0.12 (-0.13, -0.12) |
| IRS<sub>Stevenson</sub> | 0.01 (-0.04, 0.05) | 0.01 (-0.04, 0.05) |
| mdNLR | 2.10 (1.41, 2.96) | 2.12 (1.51, 2.93) |
| Estimated cell type composition |  |  |
| CD8+ T cell | 0.01 (0.00, 0.04) | 0.01 (0.00, 0.04) |
| CD4+ T cell | 0.15 (0.12, 0.20) | 0.15 (0.11, 0.19) |
| Natural killer cell | 0.06 (0.03, 0.11) | 0.06 (0.03, 0.10) |
| B cell | 0.05 (0.03, 0.07) | 0.05 (0.03, 0.07) |
| Monocyte | 0.06 (0.05, 0.09) | 0.07 (0.05, 0.09) |
| Granulocyte | 0.63 (0.54, 0.70) | 0.63 (0.56, 0.69) |
| Histology confirmed cases, n (%) |  |  |
| No |  | 110 (16.11%) |
| Yes |  | 549 (80.38%) |
| Unknown |  | 24 (3.51%) |
| Adenocarcinoma, n (%) |  |  |
| Yes |  | 393 (57.54%) |
| Other/Unknown |  | 290 (42.46%) |

Abbreviations: mdNLR = DNA methylation derived neutrophil-lymphocyte ratio
Values are median (Q1, Q3) except where indicated.

### DNAm-IRS and future lung cancer risk

Higher IRS_Ligthart_ and IRS_Wielscher_ were associated with lower risk of lung cancer across sequentially adjusted models (**Figure 2a; Table S4**). In age- and BMI-adjusted models, the HR was 0.85 (95% CI: 0.76–0.95) for IRS_Ligthart_ and 0.87 (95% CI: 0.77–0.97) for IRS_Wielscher_ per 1-SD increase. After additional adjustment for estimated leukocyte compositions, associations remained consistent (IRS_Ligthart_: HR=0.82, 95% CI: 0.72–0.93; IRS_Wielscher_: HR=0.86, 95% CI: 0.74–0.99). With further adjustment for DNAm pack-years, associations strengthened for both scores (IRS_Ligthart_: HR=0.69, 95% CI: 0.59–0.80; IRS_Wielscher_: HR=0.78, 95% CI: 0.66–0.92). No clear associations were observed for other scores. Findings were similar in analyses restricted to histology-confirmed cases (**Table S5**) and in 2-year lag analyses restricted to cases diagnosed more than 2 years after blood collection (**Table S6**). When stratified by the median interval from blood collection to diagnosis, inverse associations were more evident among cancers diagnosed more than 10.1 years after blood collection (**Table S7**).

**Figure 1.**
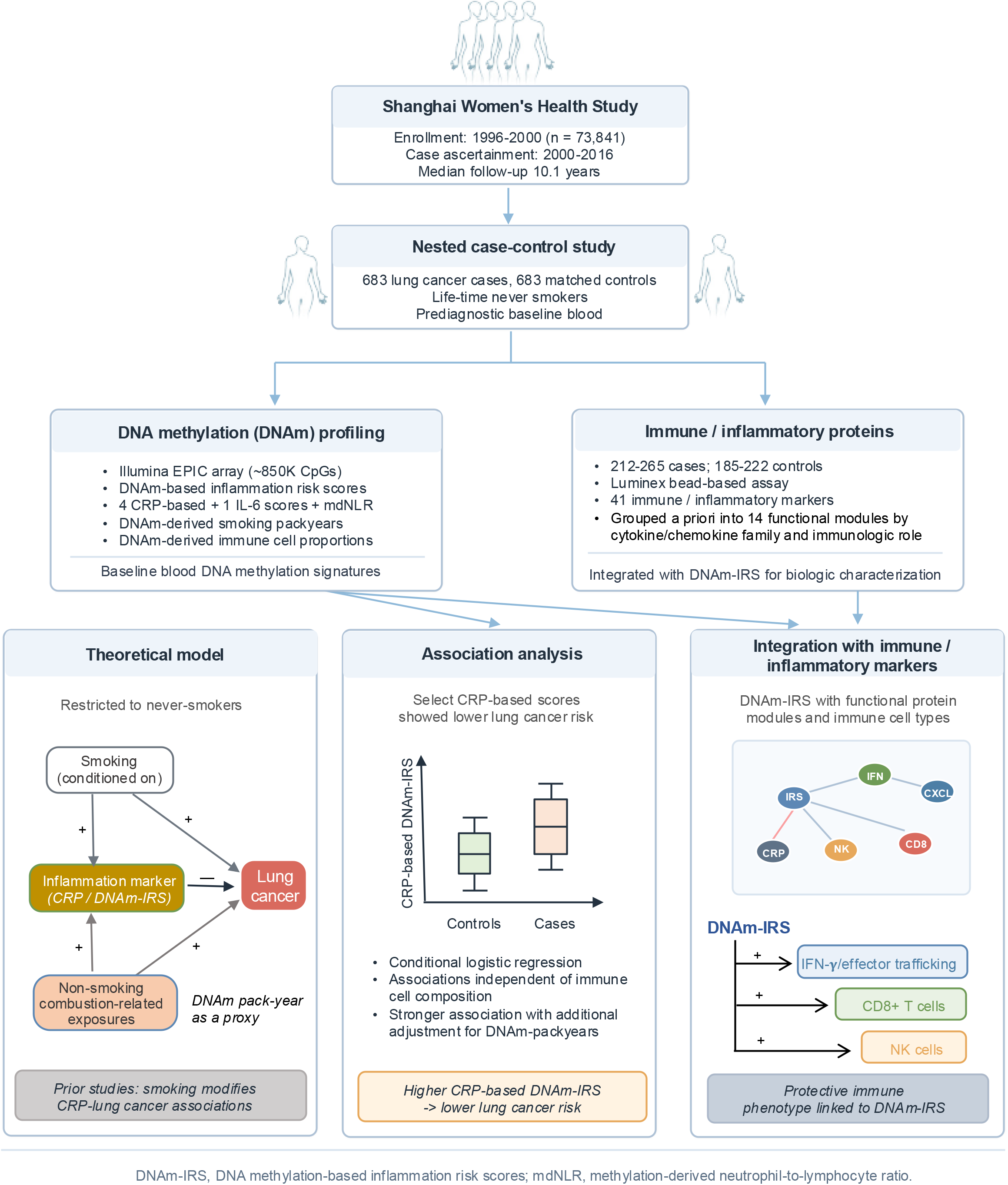
Overview of study design, molecular profiling, and analytical framework.

**Figure 2.**
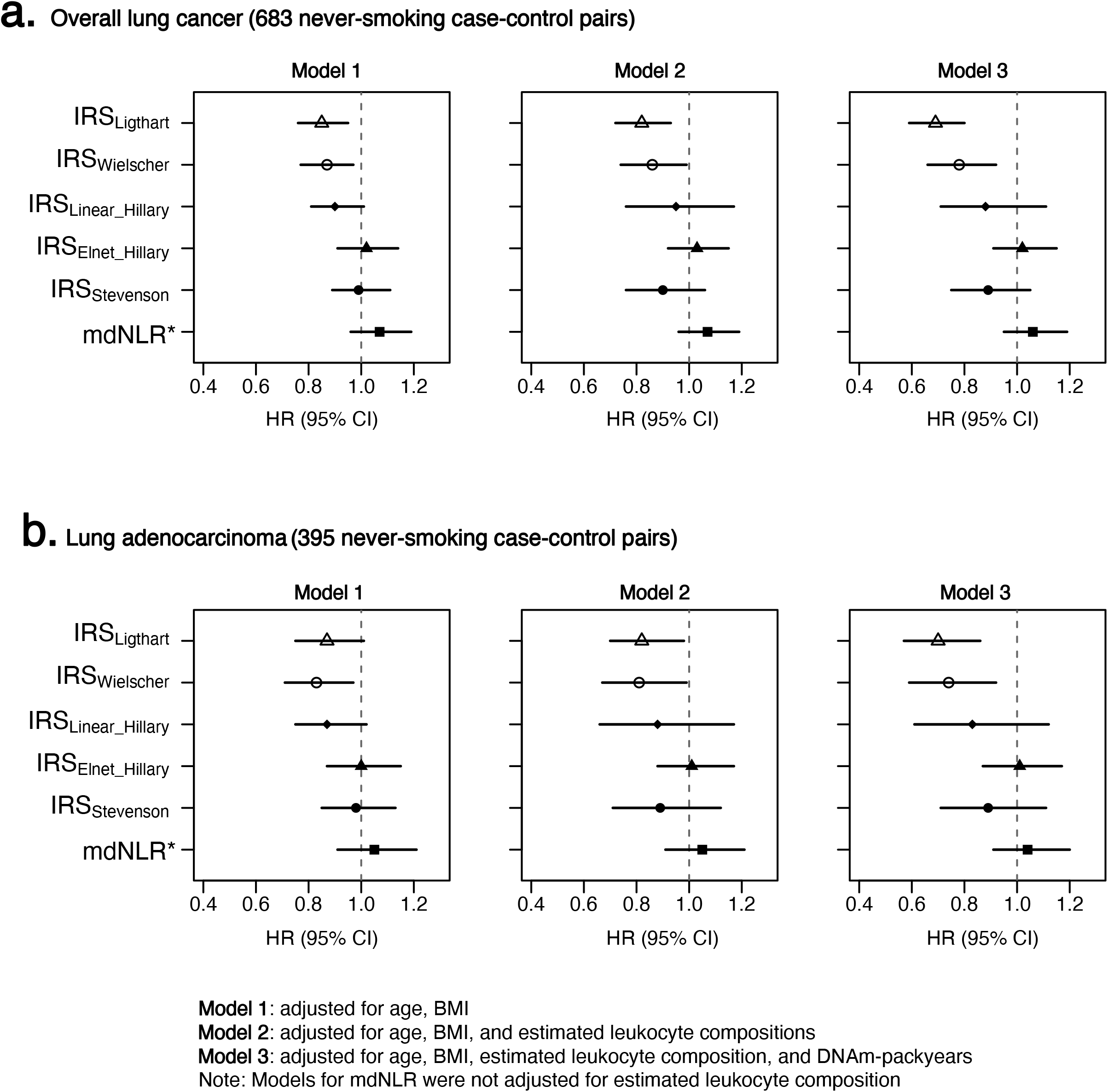
Associations of DNAm-based inflammation risk scores with lung cancer risk among never-smoking women. Hazard ratios (HRs) and 95% confidence intervals (CIs) per 1-SD increase in each DNAm-IRS are shown for (A) overall lung cancer among 683 never-smoking case-control pairs and (B) lung adenocarcinoma among 395 never-smoking case-control pairs. Estimates are presented under three models: Model 1, adjusted for age and BMI; Model 2, adjusted for age, BMI, and estimated leukocyte composition; and Model 3, adjusted for age, BMI, estimated leukocyte composition, and DNAm pack-years. DNAm-IRS include IRS_Ligthart_, IRS_Wielscher_, IRS_Linear_Hillary_, IRS_Elnet_Hillary_, IRS_Stevenson_, and methylation-derived neutrophil-to-lymphocyte ratio (mdNLR). Point estimates are indicated by symbols; horizontal lines represent 95% CIs. HRs <1 indicate lower lung cancer risk. Models for mdNLR were not adjusted for estimated leukocyte composition.

Similar patterns were observed for lung adenocarcinoma (**Figure 2b; Table S8**). In age- and BMI-adjusted models, the HR was 0.87 (95% CI: 0.75–1.01) for IRS_Ligthart_ and 0.83 (95% CI: 0.71–0.97) for IRS_Wielscher_ per 1-SD increase. These associations persisted after adjustment for estimated leukocyte compositions and strengthened after additional adjustment for DNAm pack-years (IRS_Ligthart_: HR=0.70, 95% CI: 0.57–0.86; IRS_Wielscher_: HR=0.74, 95% CI: 0.59–0.92). No clear associations were observed for the other DNAm-IRSs. Results were similar after excluding cases diagnosed within 2 years of blood collection (**Table S9**). Latency-stratified analyses also showed a pattern similar to that for overall lung cancer, although estimates were less precise because of the smaller sample size (**Table S10**).

No statistically significant interaction with DNAm pack-years was observed on the continuous scale for overall lung cancer (**Tables 2; S11**). Nevertheless, stratified analyses suggested that inverse associations for IRS_Ligthart_ and IRS_Wielscher_ were more evident among women with low DNAm pack-years stratum (IRS_Ligthart_: HR=0.65, 95% CI: 0.47-0.91 and IRS_Wielscher_: HR=0.66, 95% CI: 0.44-0.98) than in the high DNAm pack-years stratum (IRS_Ligthart_: HR=0.94, 95% CI: 0.70-1.25 and IRS_Wielscher_: HR=1.05, 95% CI: 0.76-1.47). The results were directionally similar for adenocarcinoma but less precise because of the smaller sample size (**Table S12**). By contrast, mdNLR showed a positive association in the high DNAm pack-years stratum.

**Table 2.** Associations of selected DNAm inflammation scores with overall lung cancer risk by DNAm pack-years.

|  | Overall<br>(683 pairs) | Stratified by DNAm pack-years |  | *P*-<br>*interaction* |
| --- | --- | --- | --- | --- |
|  |  | Lower<br>DNAm pack-years<br>(157 pairs) | Higher<br>DNAm pack-years<br>(180 pairs) |  |
| DNAm-IRS | HR (95% CI) | HR (95% CI) | HR (95% CI) |  |
| IRS<sub>Ligthart</sub> | **0.82 (0.71-0.95)** | **0.65 (0.47-0.91)** | 0.94 (0.70-1.25) | 0.48 |
| IRS<sub>Wielscher</sub> | **0.84 (0.71-1.00)** | **0.66 (0.44-0.98)** | 1.05 (0.76-1.47) | 0.69 |
| IRS<sub>Linear_Hillary</sub> | 0.97 (0.76-1.23) | 0.66 (0.38-1.15) | 1.23 (0.77-1.98) | 0.47 |
| Elnet_Hillary | 1.01 (0.89-1.14) | 0.96 (0.75-1.24) | 1.12 (0.89-1.42) | 0.44 |
| Stevenson | 0.95 (0.78-1.15) | 0.72 (0.49-1.05) | 1.14 (0.82-1.58) | 0.50 |
| mdNLR | 1.07 (0.95-1.21) | 1.06 (0.82-1.37) | **1.24 (1.00-1.54)** | 0.28 |

Hazard ratios (HRs) were estimated using conditional logistic regression within risk-set matched strata. Models were adjusted for age, body mass index, and estimated leukocyte proportions; mdNLR models were adjusted for age at baseline and body mass index only. P-interaction values were estimated from models including a DNAm score × DNAm pack-years product term, with DNAm pack-years modeled continuously.

### Biologic correlates of selected DNAm-IRSs

Subsequent biologic interpretation focused on IRS_Ligthart_ and IRS_Wielscher_, the two CRP-related scores associated with lung cancer risk. Both showed the most consistent associations with immune-inflammatory protein modules (**Figure 3a, Table S13**). In age- and BMI-adjusted models, higher scores were associated with higher acute-phase inflammation and IFN-γ/effector trafficking module scores; these associations were generally stronger after adjustment for estimated cell composition and for IRS_Wielscher_. Weighted partial correlations with module components showed a similar pattern (**Figure S2**).

**Figure 3.**
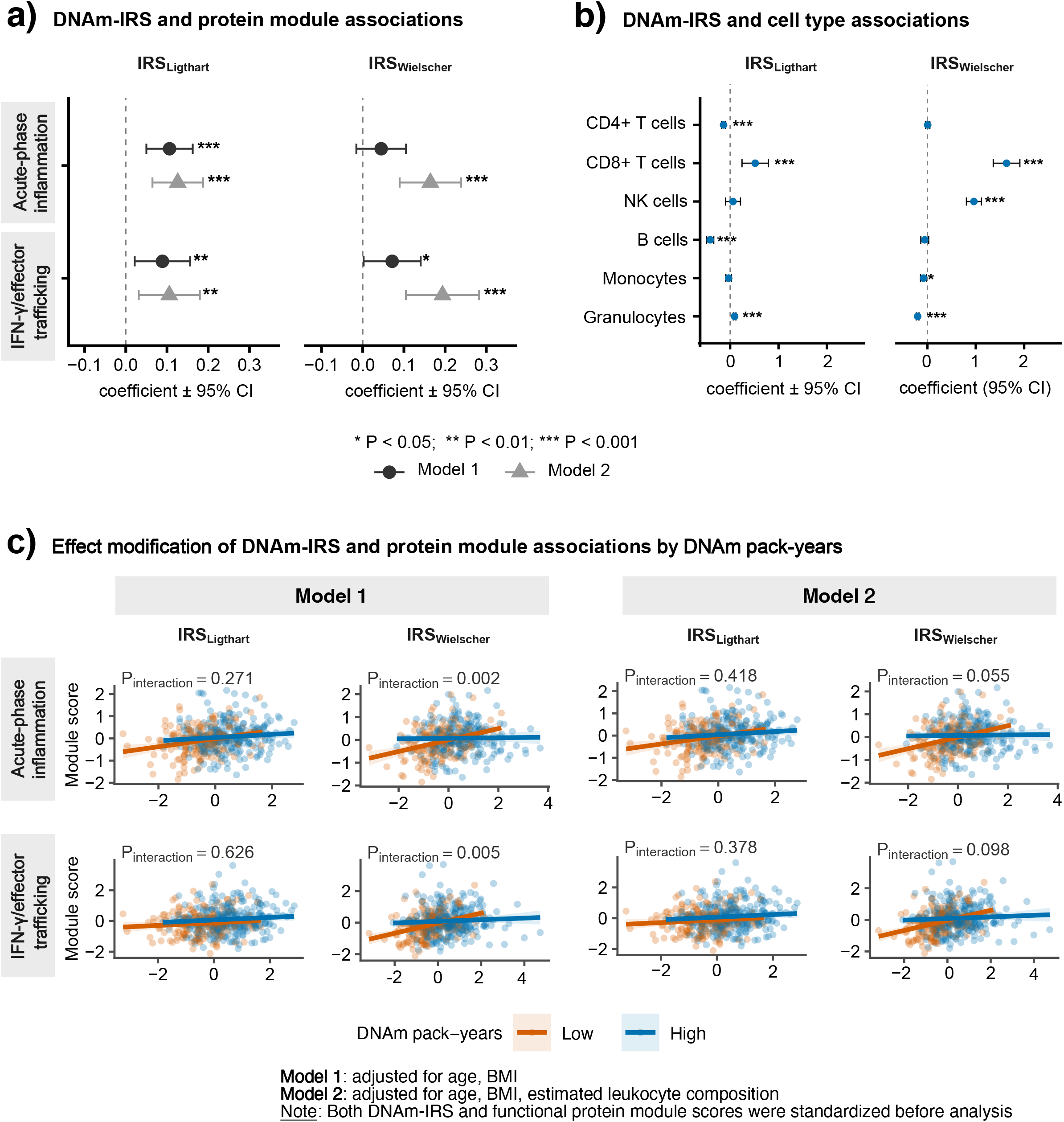
Associations of DNAm-based inflammation risk scores with selected functional immune modules and leukocyte composition, and effect modification by DNAm pack-years among never-smoking women in the SWHS. Regression coefficients (β) and 95% confidence intervals are shown for associations of IRS_Ligthart_ and IRS_Wielscher_ with (a) selected functional protein module scores and (b) DNAm-derived leukocyte composition. Estimates are presented for Model 1, adjusted for age and BMI, and Model 2, additionally adjusted for estimated leukocyte cell-type proportions where applicable. (c) Effect modification of IRS–functional protein module associations by DNAm pack-years, dichotomized at the median among controls into low and high groups. Linear fits and 95% confidence bands are shown by DNAm pack-years group under Model 1 and Model 2. Interaction P values were obtained from likelihood ratio tests. In this never-smoking population, DNAm pack-years was used as a methylation-derived proxy for cumulative smoking-related and other combustion-related exposure signals.

Associations of the targeted protein modules with lung cancer risk were generally modest and imprecise in this smaller subset (**Table S14**). Although the IFN/effector trafficking module showed an inverse association for tertile 2 versus tertile 1, this pattern was not observed for tertile 3 versus tertile 1, and the acute-phase inflammation module showed no consistent trend across tertiles. Given the suggestive effect modification by DNAm pack-years in DNAm-IRS-lung cancer associations, we next examined whether associations of IRS_Ligthart_ and IRS_Wielscher_ with these two modules varied by DNAm pack-years. DNAm pack-years modified associations of IRS_Wielscher_, but not IRS_Ligthart_, with acute-phase inflammation and IFN-γ/effector trafficking (**Figure 3c**). For IRS_Wielscher_, associations with both modules were stronger in the low DNAm pack-years stratum in the age- and BMI-adjusted model (interaction P<0.01), although this pattern was attenuated after adjustment for estimated cell composition.

The two CRP-related scores also showed distinct leukocyte composition profiles (**Figure 3b, Table S15**). Higher IRS_Ligthart_ was associated with higher CD8+ T-cell and granulocyte proportions and lower B-cell and CD4+ T-cell proportions. By contrast, higher IRS_Wielscher_ was marked by higher CD8+ T-cell and NK-cell proportions together with lower granulocyte and monocyte proportions, supporting a more cytotoxic lymphoid profile. Overall, although both scores were inversely associated with lung cancer risk, IRS_Wielscher_ appeared to capture a more specific cytotoxic immune phenotype, whereas IRS_Ligthart_ reflected a broader inflammatory-immune state. However, whether these leukocyte shifts are directly relevant to lung cancer risk remains unclear, as none of the estimated immune cell types was significantly associated with lung cancer risk after multiple-testing correction in this population (**Table S16**).

## Discussion

In this nested case-control study of never-smoking women, higher pre-diagnostic CRP-related DNAm scores, particularly IRS_Ligthart_ and IRS_Wielscher_, were associated with reduced risk of overall lung cancer and lung adenocarcinoma, the predominant histologic subtype in never smokers. These inverse associations became more evident after adjustment for DNAm pack-years, an epigenetic index that may capture broader combustion-related exposures beyond active smoking. Associations were also strengthened in longer-latency analyses, supporting that these methylation signatures may reflect longer-term biologic processes and are not driven solely by preclinical disease. In integrated analyses, these scores were associated with acute-phase inflammation and IFN-γ/effector trafficking, with stronger and more coherent patterns for IRS_Wielscher_. Our analysis further suggested a more cytotoxic lymphoid profile for IRS_Wielscher_ and a broader inflammatory-immune pattern for IRS_Ligthart_. Overall, these findings support inverse associations between selected CRP-related DNAm signatures and lung cancer risk in never-smoking women, although the relevance of these biologic correlates to risk remains uncertain and requires confirmation in studies with more comprehensive immune-inflammatory profiling.

CRP-related DNAm scores are better interpreted as epigenetic signatures of longer-term inflammatory biology than as direct surrogates for a single circulating CRP measurement, which is phasic and influenced by short-term inflammatory fluctuations. However, these signatures also appear to reflect upstream drivers of inflammation, supporting their interpretation as composite markers of chronic immune-inflammatory and exposure-related processes.[11, 14] In this cohort, we previously observed an inverse association between circulating CRP and lung cancer risk among never-smoking women,[9] a finding that might seem paradoxical given CRP’s role as a pro-inflammatory marker. The present analysis extends that observation with approximately 2.5 times more lung cancer cases and across multiple CRP-related DNAm scores, with the clearest inverse associations observed for IRS_Ligthart_ and IRS_Wielscher_. The similar inverse associations observed for more stable epigenetic inflammatory signatures strengthen the possibility that this pattern reflects longer-term host immune biology, rather than being explained solely by short-term inflammatory fluctuations. These findings are broadly consistent with inverse CRP–lung cancer associations reported among never-smokers in the Lung Cancer Cohort Consortium,[8] and with results from the smoker-prevalent CLUE I/II cohorts, in which IRS_Ligthart_ was inversely associated with lung cancer risk after rigorous adjustment for smoking using DNAm pack-years.[13]

Although all participants were self-reported never-smokers, DNAm pack-years may still capture smoking-related and other combustion-related exposure signals relevant to lung cancer risk, including household air pollution, traffic-related pollution, and secondhand smoke. Supporting this interpretation, we recently reported that among never-smoking women exposed to indoor coal combustion, DNAm pack-years positively correlated with cumulative exposure to several polycyclic aromatic hydrocarbons, including 5-methylchrysene, a potent lung carcinogen.[23] The stronger inverse associations for IRS_Ligthart_ and IRS_Wielscher_ after adjustment for DNAm pack-years therefore suggest that combustion-related methylation signals may have partially obscured their associations with lung cancer risk. Adjustment for DNAm pack-years may also reduce exposure-related methylation variation embedded within or correlated with these CRP-related scores, thereby clarifying the immune-inflammatory component of these signatures. Stratified analyses were consistent with this interpretation, with inverse associations more evident among women with lower DNAm pack-years, although interaction tests were not statistically significant. mdNLR also showed a positive association in the high DNAm pack-years stratum, a pattern broadly consistent with positive associations reported in more smoking-exposed populations. Because DNAm pack-years itself is a methylation-derived composite measure, we cannot fully exclude bias introduced by conditioning on shared exposure-related or host biologic variation. However, this is less likely to fully explain the findings given the consistency of results across adjusted and stratified analyses and the primary focus on IRS–lung cancer associations rather than on upstream determinants of these scores.

Timing is important when interpreting inflammation-related biomarkers in relation to cancer risk because such measures may reflect both longer-term host biology and changes related to occult disease. In our study, the inverse associations for the two risk-associated scores became more evident after excluding cases diagnosed within 2 years of blood collection. Consistent with this pattern, stratified analyses by median follow-up from blood collection to diagnosis (10.1 years) showed that the inverse associations were generally more evident among women with longer follow-up. Together, these patterns suggest that CRP-related DNAm scores may reflect relatively stable immune-inflammatory states with longer-term relevance to lung cancer risk, although causality cannot be inferred from these observational analyses.

A distinctive feature of this study was the evaluation of biologic correlates of DNAm-IRSs. Both IRS_Ligthart_ and IRS_Wielscher_ were positively associated with acute-phase inflammation and IFN-γ/effector trafficking, with generally stronger and more coherent patterns observed for IRS_Wielscher_, particularly after adjustment for estimated leukocyte composition. These IRS–protein module associations also appeared to vary by DNAm pack-years, suggesting that combustion-related methylation burden may influence the immune-inflammatory signals captured by these scores. The IFN-γ–inducible CXCL9/CXCL10/CXCL11–CXCR3 axis is implicated in cytotoxic effector-cell trafficking and antitumor immunity.[35] Although this supports the biologic plausibility of an immune-surveillance phenotype captured by DNAm-IRSs, the IFN-γ/effector-trafficking module showed only modest, nonmonotonic inverse association with lung cancer risk, and circulating levels of its component markers were not independently associated with risk in this population.[9]

Leukocyte analyses likewise suggested distinct profiles, with **I**RS_Wielscher_ characterized by higher estimated CD8+ T cells and NK cells and lower granulocyte and monocyte estimates, consistent with a more cytotoxic lymphoid profile, whereas **I**RS_Ligthart_ appeared to reflect a broader inflammatory-immune pattern. Although no estimated immune cell type was significantly associated with lung cancer risk in our study, the IRS_Wielscher_-associated profile is directionally consistent with prior prospective studies linking more myeloid-skewed profiles, including higher mdNLR and higher monocyte- or neutrophil-plus-monocyte-to-lymphocyte ratios, to increased lung cancer risk.[13] It is also consistent with findings from the EPIC-Heidelberg cohort in which higher relative CD8□ T-cell proportions were associated with lower lung cancer risk.[36] Importantly, IRS–lung cancer associations persisted after adjustment for estimated leukocyte composition, suggesting that they were not explained solely by circulating immune-cell proportions.

This study has several strengths. The prospective design with pre-diagnostic biospecimens reduces concern about reverse causation, and latency analyses further supported the temporal robustness of the observed associations. Restriction to never-smoking women limits confounding by active smoking, while adjustment for DNAm pack-years helps address residual exposure measurement error and broader combustion-related inflammatory exposures. The relatively large sample size for lung cancer in never-smokers, together with concordant inverse associations across multiple independently derived CRP-related DNAm-IRS algorithms, supports the robustness of the findings. Integration of DNAm-IRS with circulating immune-inflammatory proteins and DNAm-derived leukocyte composition enabled systematic characterization of the broader immune-inflammatory signatures captured by these scores. Concordant inverse associations across multiple independently derived CRP-related DNAm scores, together with consistency with findings from the smoker-prevalent CLUE I/II cohorts after rigorous adjustment for smoking using DNAm pack-years,[13] support robustness and potential generalizability.

Several limitations should be considered. First, although this study included a relatively large number of lung cancer cases among never-smokers, power was limited for stratified analyses, formal interaction testing, and histology-specific analyses beyond adenocarcinoma. Second, most DNAm-IRS algorithms were developed in predominantly European-ancestry populations, and validation of DNAm-IRS–lung cancer associations in non-European populations remains limited. Nonetheless, the concordance between our findings in East Asian never-smoking women and those from the predominantly White, smoker-prevalent CLUE I/II cohorts provides partial reassurance. Third, DNAm-IRSs were constructed using available post-QC CpG sites, and some CpG sites from the original algorithms were not retained after methylation quality control, which may have introduced measurement error and reduced comparability with the originally developed scores. Fourth, leukocyte composition was estimated from DNAm rather than measured directly, and circulating immune proteins were assessed at a single pre-diagnostic time point. Fifth, the immune-inflammatory marker panel was limited; broader circulating protein panels will be needed to more comprehensively define the immune pathways captured by DNAm-IRSs and their relevance to lung cancer risk. Finally, generalizability to men, smokers, and other racial or ethnic populations requires further evaluation.

In conclusion, higher pre-diagnostic CRP-related DNAm scores, particularly IRS_Ligthart_ and IRS_Wielscher_, were associated with lower lung cancer risk among never-smoking women. These associations were more evident after adjustment for DNAm pack-years and in longer-latency analyses, supporting the possibility that selected CRP-related methylation signatures capture relatively stable immune-inflammatory phenotypes relevant to lung cancer susceptibility. Integrated protein and leukocyte analyses further suggest that these scores reflect broader immune-inflammatory and exposure-related biology, including acute-phase inflammation, IFN-γ/effector trafficking, and cytotoxic lymphoid features. Further studies in diverse populations with detailed exposure assessment and broader immune profiling are needed to validate these findings and clarify the biologic pathways linking CRP-related DNAm signatures to lung cancer risk. More broadly, these findings suggest that immune-inflammatory biomarkers may not have uniform associations with lung cancer risk across exposure settings, underscoring the need to consider exposure context in future etiologic and biomarker studies.

## Data Availability

All data produced in the present work are contained in the manuscript

## Abbreviations

BMI: Body Mass Index
CGR: Cancer Genomics Research Laboratory
CLUE: Campaigns for Lung Cancer Prevention and Early Detection in the United States (I/II cohorts)
CRP: C-reactive protein
CXCR3: C-X-C motif chemokine receptor 3
DNAm: DNA methylation
FDR: False discovery rate
HR: Hazard ratio
ICD-9: International Classification of Diseases, Ninth Revision
ICD-O-2: International Classification of Diseases for Oncology, Second Edition
IL: Interleukin
IRS: Inflammation risk score
mdNLR: Methylation-derived neutrophil-to-lymphocyte ratio
NK: Natural killer cells
PAH: Polycyclic aromatic hydrocarbon
SD: Standard deviation
SWHS: Shanghai Women’s Health Study

## Acknowledgements

The authors acknowledge the research contributions of the Cancer Genomics Research Laboratory of the Intramural Research Program, National Cancer Institute, National Institutes of Health for their expertise, execution, and support of this research in the areas of project planning, wet laboratory processing of specimens, and generating the data.

## Ethics approval and consent to participate

All study participants provided written informed consent before being interviewed, and the study protocols were approved by the institutional review boards of all participating institutions (number OH98CN006).

## Submission declaration

All authors have approved this submission and the responsible authorities where the work was conducted have no objection to its publication. The work has not been published previously, is not under consideration elsewhere, and will not be published in the same form in any language, including electronically, without written permission from the copyright holder if accepted.

## Authorship

N.R., Q.L., W.Z., J.Y.Y.W., and X.O.S. designed the study and oversaw data collection. M.L.R. led the intellectual and analytical work of the study, including formulating the research question, developing the analysis plan, performing the analyses, interpreting the results, and writing the manuscript. A.G. contributed to data analysis. All authors contributed to the interpretation of the findings, critically revised the manuscript for important intellectual content, and approved the final version. M.L.R. and Q.L. served as guarantors of the work, with full access to all study data, and take responsibility for the integrity of the data and the accuracy of the analyses. The work reported in this paper was performed by the authors, unless otherwise specified in the text.

## Declaration of Generative AI and AI-assisted technologies in the writing process

During the preparation of this work the author used ChatGPT to improve readability and language. After using this tool, the author reviewed and edited the content as needed and takes full responsibility for the content of the publication.

## References

1. Sun, S., J.H. Schiller, and A.F. Gazdar, Lung cancer in never smokers--a different disease. Nat Rev Cancer, 2007. 7(10): p. 778–90.

2. Murphy, C., T. Pandya, C. Swanton, and B.J. Solomon, Lung Cancer in Nonsmoking Individuals: A Review. JAMA, 2025. 334(20): p. 1836–1845.

3. Hanahan, D. and Robert A. Weinberg, Hallmarks of Cancer: The Next Generation. Cell, 2011. 144(5): p. 646–674.

4. Zhao, H., L. Wu, G. Yan, Y. Chen, M. Zhou, Y. Wu, and Y. Li, Inflammation and tumor progression: signaling pathways and targeted intervention. Signal Transduction and Targeted Therapy, 2021. 6(1): p. 263.

5. Shiels, M.S., H.A. Katki, A. Hildesheim, R.M. Pfeiffer, E.A. Engels, M. Williams, T.J. Kemp, N.E. Caporaso, L.A. Pinto, and A.K. Chaturvedi, Circulating Inflammation Markers, Risk of Lung Cancer, and Utility for Risk Stratification. J Natl Cancer Inst, 2015. 107(10).

6. Shiels, M.S., R.M. Pfeiffer, A. Hildesheim, E.A. Engels, T.J. Kemp, J.-H. Park, H.A. Katki, J. Koshiol, G. Shelton, N.E. Caporaso, L.A. Pinto, and A.K. Chaturvedi, Circulating Inflammation Markers and Prospective Risk for Lung Cancer. JNCI: Journal of the National Cancer Institute, 2013. 105(24): p. 1871–1880.

7. Chaturvedi, A.K., N.E. Caporaso, H.A. Katki, H.L. Wong, N. Chatterjee, S.R. Pine, S.J. Chanock, J.J. Goedert, and E.A. Engels, C-reactive protein and risk of lung cancer. J Clin Oncol, 2010. 28(16): p. 2719–26.

8. Muller, D.C., T.L. Larose, A. Hodge, F. Guida, A. Langhammer, K. Grankvist, K. Meyer, Q. Cai, A.A. Arslan, A. Zeleniuch-Jacquotte, D. Albanes, G.G. Giles, H.D. Sesso, I.M. Lee, J.M. Gaziano, J.M. Yuan, J. Hoffman Bolton, J.E. Buring, K. Visvanathan, L. Le Marchand, M.P. Purdue, N.E. Caporaso, Ø. Midttun, P.M. Ueland, R.L. Prentice, S.J. Weinstein, V.L. Stevens, W. Zheng, W.J. Blot, X.O. Shu, X. Zhang, Y.B. Xiang, W.P. Koh, K. Hveem, C.A. Thomson, M. Pettinger, G. Engström, H. Brunnström, R.L. Milne, M.J. Stampfer, J. Han, M. Johansson, P. Brennan, G. Severi, and M. Johansson, Circulating high sensitivity C reactive protein concentrations and risk of lung cancer: nested case-control study within Lung Cancer Cohort Consortium. Bmj, 2019. 364: p. k4981.

9. Shiels, M.S., X.O. Shu, A.K. Chaturvedi, Y.T. Gao, Y.B. Xiang, Q. Cai, W. Hu, G. Shelton, B.T. Ji, L.A. Pinto, T.J. Kemp, N. Rothman, W. Zheng, A. Hildesheim, and Q. Lan, A prospective study of immune and inflammation markers and risk of lung cancer among female never smokers in Shanghai. Carcinogenesis, 2017. 38(10): p. 1004–1010.

10. Pepys, M.B. and G.M. Hirschfield, C-reactive protein: a critical update. J Clin Invest, 2003. 111(12): p. 1805–12.

11. Ligthart, S., C. Marzi, S. Aslibekyan, M.M. Mendelson, K.N. Conneely, T. Tanaka, E. Colicino, L.L. Waite, R. Joehanes, W. Guan, J.A. Brody, C. Elks, R. Marioni, M.A. Jhun, G. Agha, J. Bressler, C.K. Ward-Caviness, B.H. Chen, T. Huan, K. Bakulski, E.L. Salfati, G. Fiorito, S. Wahl, K. Schramm, J. Sha, D.G. Hernandez, A.C. Just, J.A. Smith, N. Sotoodehnia, L.C. Pilling, J.S. Pankow, P.S. Tsao, C. Liu, W. Zhao, S. Guarrera, V.J. Michopoulos, A.K. Smith, M.J. Peters, D. Melzer, P. Vokonas, M. Fornage, H. Prokisch, J.C. Bis, A.Y. Chu, C. Herder, H. Grallert, C. Yao, S. Shah, A.F. McRae, H. Lin, S. Horvath, D. Fallin, A. Hofman, N.J. Wareham, K.L. Wiggins, A.P. Feinberg, J.M. Starr, P.M. Visscher, J.M. Murabito, S.L. Kardia, D.M. Absher, E.B. Binder, A.B. Singleton, S. Bandinelli, A. Peters, M. Waldenberger, G. Matullo, J.D. Schwartz, E.W. Demerath, A.G. Uitterlinden, J.B. van Meurs, O.H. Franco, Y.I. Chen, D. Levy, S.T. Turner, I.J. Deary, K.J. Ressler, J. Dupuis, L. Ferrucci, K.K. Ong, T.L. Assimes, E. Boerwinkle, W. Koenig, D.K. Arnett, A.A. Baccarelli, E.J. Benjamin, and A. Dehghan, DNA methylation signatures of chronic lowgrade inflammation are associated with complex diseases. Genome Biol, 2016. 17(1): p. 255.

12. Hillary, R.F., H.K. Ng, D.L. McCartney, H.R. Elliott, R.M. Walker, A. Campbell, F. Huang, K. Direk, P. Welsh, N. Sattar, J. Corley, C. Hayward, A.M. McIntosh, C. Sudlow, K.L. Evans, S.R. Cox, J.C. Chambers, M. Loh, C.L. Relton, R.E. Marioni, P.D. Yousefi, and M. Suderman, Blood-based epigenome-wide analyses of chronic low-grade inflammation across diverse population cohorts. Cell Genom, 2024. 4(5): p. 100544.

13. Zhao, N., M. Ruan, D.C. Koestler, J. Lu, L.A. Salas, K.T. Kelsey, E.A. Platz, and D.S. Michaud, Methylation-derived inflammatory measures and lung cancer risk and survival. Clinical Epigenetics, 2021. 13(1): p. 222.

14. Wielscher, M., P.R. Mandaviya, B. Kuehnel, R. Joehanes, R. Mustafa, O. Robinson, Y. Zhang, B. Bodinier, E. Walton, P.P. Mishra, P. Schlosser, R. Wilson, P.-C. Tsai, S. Palaniswamy, R.E. Marioni, G. Fiorito, G. Cugliari, V. Karhunen, M. Ghanbari, B.M. Psaty, M. Loh, J.C. Bis, B. Lehne, N. Sotoodehnia, I.J. Deary, M. Chadeau-Hyam, J.A. Brody, A. Cardona, E. Selvin, A.K. Smith, A.H. Miller, M.A. Torres, E. Marouli, X. Gào, J.B.J. van Meurs, J. Graf-Schindler, W. Rathmann, W. Koenig, A. Peters, W. Weninger, M. Farlik, T. Zhang, W. Chen, Y. Xia, A. Teumer, M. Nauck, H.J. Grabe, M. Doerr, T. Lehtimäki, W. Guan, L. Milani, T. Tanaka, K. Fisher, L.L. Waite, S. Kasela, P. Vineis, N. Verweij, P. van der Harst, L. Iacoviello, C. Sacerdote, S. Panico, V. Krogh, R. Tumino, E. Tzala, G. Matullo, M.A. Hurme, O.T. Raitakari, E. Colicino, A.A. Baccarelli, M. Kähönen, K.-H. Herzig, S. Li, K.N. Conneely, J.S. Kooner, A. Köttgen, B.T. Heijmans, P. Deloukas, C. Relton, K.K. Ong, J.T. Bell, E. Boerwinkle, P. Elliott, H. Brenner, M. Beekman, D. Levy, M. Waldenberger, J.C. Chambers, A. Dehghan, M.-R. Järvelin, and B. consortium, DNA methylation signature of chronic low-grade inflammation and its role in cardio-respiratory diseases. Nature Communications, 2022. 13(1): p. 2408.

15. Koestler, D.C., J. Usset, B.C. Christensen, C.J. Marsit, M.R. Karagas, K.T. Kelsey, and J.K. Wiencke, DNA Methylation-Derived Neutrophil-to-Lymphocyte Ratio: An Epigenetic Tool to Explore Cancer Inflammation and Outcomes. Cancer Epidemiology, Biomarkers & Prevention, 2017. 26(3): p. 328–338.

16. Stevenson, A.J., D.A. Gadd, R.F. Hillary, D.L. McCartney, A. Campbell, R.M. Walker, K.L. Evans, S.E. Harris, T.L. Spires-Jones, A.F. McRae, P.M. Visscher, A.M. McIntosh, I.J. Deary, and R.E. Marioni, Creating and Validating a DNA Methylation-Based Proxy for Interleukin-6. The Journals of Gerontology: Series A, 2021. 76(12): p. 2284–2292.

17. Pine, S.R., L.E. Mechanic, L. Enewold, A.K. Chaturvedi, H.A. Katki, Y.L. Zheng, E.D. Bowman, E.A. Engels, N.E. Caporaso, and C.C. Harris, Increased levels of circulating interleukin 6, interleukin 8, C-reactive protein, and risk of lung cancer. J Natl Cancer Inst, 2011. 103(14): p. 1112–22.

18. Brenner, D.R., A. Fanidi, K. Grankvist, D.C. Muller, P. Brennan, J. Manjer, G. Byrnes, A. Hodge, G. Severi, G.G. Giles, M. Johansson, and M. Johansson, Inflammatory Cytokines and Lung Cancer Risk in 3 Prospective Studies. Am J Epidemiol, 2017. 185(2): p. 86–95.

19. Nøst, T.H., K. Alcala, I. Urbarova, K.S. Byrne, F. Guida, T.M. Sandanger, and M. Johansson, Systemic inflammation markers and cancer incidence in the UK Biobank. Eur J Epidemiol, 2021. 36(8): p. 841–848.

20. Joehanes, R., A.C. Just, R.E. Marioni, L.C. Pilling, L.M. Reynolds, P.R. Mandaviya, W. Guan, T. Xu, C.E. Elks, S. Aslibekyan, H. Moreno-Macias, J.A. Smith, J.A. Brody, R. Dhingra, P. Yousefi, J.S. Pankow, S. Kunze, S.H. Shah, A.F. McRae, K. Lohman, J. Sha, D.M. Absher, L. Ferrucci, W. Zhao, E.W. Demerath, J. Bressler, M.L. Grove, T. Huan, C. Liu, M.M. Mendelson, C. Yao, D.P. Kiel, A. Peters, R. Wang-Sattler, P.M. Visscher, N.R. Wray, J.M. Starr, J. Ding, C.J. Rodriguez, N.J. Wareham, M.R. Irvin, D. Zhi, M. Barrdahl, P. Vineis, S. Ambatipudi, A.G. Uitterlinden, A. Hofman, J. Schwartz, E. Colicino, L. Hou, P.S. Vokonas, D.G. Hernandez, A.B. Singleton, S. Bandinelli, S.T. Turner, E.B. Ware, A.K. Smith, T. Klengel, E.B. Binder, B.M. Psaty, K.D. Taylor, S.A. Gharib, B.R. Swenson, L. Liang, D.L. DeMeo, G.T. O’Connor, Z. Herceg, K.J. Ressler, K.N. Conneely, N. Sotoodehnia, S.L. Kardia, D. Melzer, A.A. Baccarelli, J.B. van Meurs, I. Romieu, D.K. Arnett, K.K. Ong, Y. Liu, M. Waldenberger, I.J. Deary, M. Fornage, D. Levy, and S.J. London, Epigenetic Signatures of Cigarette Smoking. Circ Cardiovasc Genet, 2016. 9(5): p. 436–447.

21. Yanbaeva, D.G., M.A. Dentener, E.C. Creutzberg, G. Wesseling, and E.F.M. Wouters, Systemic Effects of Smoking. Chest, 2007. 131(5): p. 1557–1566.

22. Hecht, S.S., Tobacco Smoke Carcinogens and Lung Cancer. JNCI: Journal of the National Cancer Institute, 1999. 91(14): p. 1194–1210.

23. Rahman, M.L., L. Portengen, B. Blechter, C.E. Breeze, J.Y.Y. Wong, W. Hu, G.S. Downward, Y. Zhang, A. Cardenas, B. Ning, J. Li, K. Yang, H.D. Hosgood, D.T. Silverman, N. Rothman, Y. Huang, R. Vermeulen, and Q. Lan, Epigenome-wide association study of household air pollution exposure in an area with high lung cancer incidence. Environmental Research, 2026. 292: p. 123690.

24. Blechter, B., A. Cardenas, J. Shi, J.Y.Y. Wong, W. Hu, M.L. Rahman, C. Breeze, G.S. Downward, L. Portengen, Y. Zhang, B. Ning, B.T. Ji, R. Cawthon, J. Li, K. Yang, A. Bozack, H. Dean Hosgood, D.T. Silverman, Y. Huang, N. Rothman, R. Vermeulen, and Q. Lan, Household air pollution and epigenetic aging in Xuanwei, China. Environ Int, 2023. 178: p. 108041.

25. Lu, A.T., A. Quach, J.G. Wilson, A.P. Reiner, A. Aviv, K. Raj, L. Hou, A.A. Baccarelli, Y. Li, J.D. Stewart, E.A. Whitsel, T.L. Assimes, L. Ferrucci, and S. Horvath, DNA methylation GrimAge strongly predicts lifespan and healthspan. Aging, 2019. 11(2): p. 303–327.

26. Yao, Y., K. Wolf, S. Breitner, S. Zhang, M. Waldenberger, J. Winkelmann, A. Schneider, and A. Peters, Long-term exposure to traffic-related air pollution is associated with epigenetic age acceleration. Environmental Research, 2026. 288: p. 123284.

27. Tantoh, D.M., M.-C. Wu, C.-C. Chuang, P.-H. Chen, Y.S. Tyan, O.N. Nfor, W.-Y. Lu, and Y.-P. Liaw, AHRR cg05575921 methylation in relation to smoking and PM2.5 exposure among Taiwanese men and women. Clinical Epigenetics, 2020. 12(1): p. 117.

28. Wu, Y., R. Xu, S. Li, B. Wen, M.C. Southey, P.-A. Dugue, J.L. Hopper, M.J. Abramson, S. Li, and Y. Guo, Association between wildfire-related PM2.5 and epigenetic aging: A twin and family study in Australia. Journal of Hazardous Materials, 2025. 481: p. 136486.

29. Zheng, W., W.H. Chow, G. Yang, F. Jin, N. Rothman, A. Blair, H.L. Li, W. Wen, B.T. Ji, Q. Li, X.O. Shu, and Y.T. Gao, The Shanghai Women’s Health Study: rationale, study design, and baseline characteristics. Am J Epidemiol, 2005. 162(11): p. 1123–31.

30. Tian, Y., T.J. Morris, A.P. Webster, Z. Yang, S. Beck, A. Feber, and A.E. Teschendorff, ChAMP: updated methylation analysis pipeline for Illumina BeadChips. Bioinformatics, 2017. 33(24): p. 3982–3984.

31. Cover, T. and P. Hart, Nearest neighbor pattern classification. IEEE Transactions on Information Theory, 1967. 13(1): p. 21–27.

32. Teschendorff, A.E., F. Marabita, M. Lechner, T. Bartlett, J. Tegner, D. Gomez-Cabrero, and S. Beck, A beta-mixture quantile normalization method for correcting probe design bias in Illumina Infinium 450 k DNA methylation data. Bioinformatics, 2013. 29(2): p. 189–96.

33. Johnson, W.E., C. Li, and A. Rabinovic, Adjusting batch effects in microarray expression data using empirical Bayes methods. Biostatistics, 2007. 8(1): p. 118–27.

34. Houseman, E.A., W.P. Accomando, D.C. Koestler, B.C. Christensen, C.J. Marsit, H.H. Nelson, J.K. Wiencke, and K.T. Kelsey, DNA methylation arrays as surrogate measures of cell mixture distribution. BMC Bioinformatics, 2012. 13(1): p. 86.

35. Tokunaga, R., W. Zhang, M. Naseem, A. Puccini, M.D. Berger, S. Soni, M. McSkane, H. Baba, and H.J. Lenz, CXCL9, CXCL10, CXCL11/CXCR3 axis for immune activation - A target for novel cancer therapy. Cancer Treat Rev, 2018. 63: p. 40–47.

36. Le Cornet, C., K. Schildknecht, A. Rossello Chornet, R.T. Fortner, S. González Maldonado, V.A. Katzke, T. Kühn, T. Johnson, S. Olek, and R. Kaaks, Circulating Immune Cell Composition and Cancer Risk: A Prospective Study Using Epigenetic Cell Count Measures. Cancer Res, 2020. 80(9): p. 1885–1892.

