## Supplemental information for "Epigenetic inflammation signatures and lung cancer risk among never-smoking women: a nested case-control study"

**Material and Methods**

***Study population and design***

This nested case-control study was conducted within the Shanghai Women’s Health Study (SWHS), a population-based prospective cohort of 74,941 women enrolled in Shanghai, China, from 1997 to 2000, as previously described.[1] At enrollment and during follow-up, participants completed structured in-person interviews that collected information on demographic characteristics, occupational and environmental exposures, lifestyle factors, and diet. The participation rate in the parent cohort was 92.7%.

Incident cancers were identified through active follow-up surveys conducted every 2-3 years and by annual linkage with the Shanghai Cancer Registry and the Vital Statistics Unit. Eligible cases were women with incident lung cancer diagnosed during follow-up who were lifetime never-smokers, defined according to the U.S. Centers for Disease Control and Prevention criterion as having smoked fewer than 100 cigarettes in their lifetime. For each case, matched controls were selected using risk-set sampling (incidence density sampling) from cohort members who were alive and cancer free at the time of the case diagnosis, with matching on date of birth (±2 years) and date of blood collection (±3 months).

Lung cancers were identified using ICD-9 code 162. Histology was classified using ICD-O-2 morphology codes as adenocarcinoma, non-adenocarcinoma, or unclassified (80003, 80413, 80703, 81403, 82403, 82603, 84803, 85503, 85603). Cases were diagnosed between 2000 and 2014, with a median follow-up of 10.1 years (range, 0.1-17.3 years). Controls were individually matched to cases on date of birth (±2 years) and date of biospecimen collection (±6 months). Overall, 1,372 participants (686 matched case-control pairs) were selected.

All participants provided written informed consent. Study protocols were approved by the institutional review boards of all participating institutions.

***DNA extraction and DNA methylation measurement***

Genomic DNA was extracted from buffy coat at baseline and processed for DNA methylation profiling at the NCI Cancer Genomics Research Laboratory using standardized laboratory procedures. Briefly, 400 ng of DNA per sample was quantified using the Quant-iT PicoGreen dsDNA assay (Thermo Fisher Scientific, Waltham, MA) and sodium bisulfite-converted using the EZ-96 DNA Methylation MagPrep Kit (Zymo Research, Irvine, CA), according to the manufacturer’s instructions. This procedure converts unmethylated cytosines to uracil while leaving 5-methylcytosine and 5-hydroxymethylcytosine unchanged. An internal control sample (NA07057; Coriell Cell Repositories, Camden, NJ) was included every 95 samples to monitor bisulfite conversion performance. Bisulfite-converted DNA was hybridized to the Illumina Infinium MethylationEPIC BeadChip, which interrogates approximately 850,000 CpG sites.

***Methylation preprocessing and quality control***

Raw DNA methylation IDAT files were processed in R ([www.r-project.org/](http://www.r-project.org/)) using Bioconductor packages,[2] implementing the ChAMP pipeline with default settings.[3] Probe-level filtering removed 66,597 probes with detection p-value >0.01, 1,061 probes with bead count <3 in at least 5% of samples, 2,449 non-CpG probes, 87,808 SNP-annotated probes, 11 multi-mapping probes, and 15,710 sex chromosome probes, leaving 692,282 probes for analysis. Missing values were imputed using the K-nearest neighbor method [4] and Beta Mixture Quantile dilation (BMIQ) was used to normalize beta values [5]. Batch effects related to array processing (plate, array, slide) were corrected using ComBat.[6] We estimated cell-type fractions from normalized EPIC beta values using the Houseman approach.[7] Of the 686 case-control pairs initially selected, two control samples were excluded during DNA extraction/laboratory processing and one additional control sample failed during IDAT file import; the final analytical set comprised 686 cases and 683 controls.

***Calculation of DNAm-based IRS indices***

DNAm-based IRS were calculated using published algorithms, restricting to CpG sites available in our dataset after quality control, ComBat batch correction, and probe filtering. IRS_Ligthart_ was computed using 52 of 58 CRP-associated CpGs reported by Ligthart et al. (2016).[8] IRS_Wielscher_ was computed using 1,333 of 1,765 CRP-associated CpGs reported by Wielscher et al. (2022).[9] IRS_Linear_Hillary_ and IRS_Elnet_Hillary_ were derived using 32,196 of 33,939 and 1,334 of 1,467 CpGs, respectively, from Hillary et al., who reported cross-cohort generalizability of these indices, including across ancestry groups.[10] IRS_Stevenson_ was computed using 34 of 35 IL-6–associated CpGs reported by Stevenson et al. (2021).[11] Finally, mdNLR was derived from DNAm-predicted leukocyte proportions as the granulocyte-to-lymphocyte ratio, applying a minimum lymphocyte proportion of 0.01 for numerical stability, and was log-transformed. All IRS indices were standardized prior to analysis. DNAm pack-years was calculated as a weighted sum of smoking-associated CpG sites (1,580 CpGs) using published regression coefficients for pack-years as weights.[12]

***Circulating immune–inflammatory markers***

A total of 61 immune–inflammatory markers were quantified using Luminex bead-based assays (Millipore, Billerica, MA), as previously described.[13] Markers meeting pre-specified analytic criteria (≥90% detectable, within-batch CV ≤15%, and intraclass correlation coefficient ≥0.80) were grouped a priori into functional modules based on cytokine/chemokine families and canonical immunologic roles (Table S1). Protein concentrations were log-transformed (adding a small constant to avoid log[0]) and standardized; module scores were calculated as the within-sample mean of z-scored marker values within each module. The revised modules comprised acute-phase/systemic inflammation (CRP, SAA, SAP, IL-6, TNF-α); type 1 IFN effector trafficking (CXCL9, CXCL10, CXCL11); myeloid innate recruitment (MCP-1, MCP-2, MCP-4, MIP-1α, MIP-1β, MIP-1δ); neutrophilic chemotaxis (GRO, ENA-78, CXCL6, IL-8); lymphoid organization/homing (CCL19, CCL21, CXCL13, CXCL12); type 2/barrier chemotaxis (Eotaxin, Eotaxin-2, TARC, MDC, CTACK); Th17/mucosal inflammation (IL-17A, IL-23, CCL20); immune counter-regulation (IL-10, IL-1RA); immune activation cytokines (IL-12p70, GM-CSF, IL-1β, IL-21); and five single-marker modules representing type 2 cytokine signaling (IL-13), effector cell trafficking (Fractalkine), lymphocyte chemoattraction (IL-16), lymphocyte homeostasis (IL-7), and cytotoxic/apoptosis signaling (TRAIL).

**Statistical analysis**

To provide biological context for the DNAm-IRS associations with lung cancer, we examined the extent to which selected DNAm-IRS were related to circulating immune-inflammatory proteins, prespecified functional protein module scores, and estimated leukocyte composition. Associations between selected DNAm-IRS and protein module scores were evaluated using linear regression adjusted for age at baseline and body mass index (BMI), with additional adjustment for estimated leukocyte proportions in a second model. Associations between DNAm-IRS and estimated leukocyte proportions were similarly assessed using linear regression adjusted for age and BMI, with cell proportions logit-transformed before analysis. To evaluate the relevance of these immune-inflammatory protein modules to lung cancer risk, module scores were categorized into tertiles based on the control distribution, and associations with lung cancer and adenocarcinoma were estimated using conditional logistic regression with the lowest tertile as the reference group. We also evaluated associations of estimated immune cell proportions, modeled continuously, with lung cancer risk using conditional logistic regression. To further contextualize these findings, we estimated partial Spearman correlations between DNAm-IRS and immune-inflammatory proteins and functional module scores, adjusting for age and BMI in the base model and additionally for DNAm-estimated leukocyte proportions in the extended model. Pairwise correlations among protein module scores were evaluated using Pearson correlation coefficients.

To examine whether DNAm-estimated smoking pack-years modified the association between each IRS and immune module scores, we dichotomized participants into low and high DNAm pack-years groups using the median value derived from control participants. For each IRS–immune module pair, we fitted two linear regression models — one with main effects only and one including an IRS × DNAm pack-years interaction term — and compared them by likelihood ratio test. Interaction p-values are reported for each IRS across all 14 immune modules. Associations were visualized as stratified scatter plots with linear fits and 95% confidence bands.

**References**

1.	Zheng, W., W.H. Chow, G. Yang, F. Jin, N. Rothman, A. Blair, H.L. Li, W. Wen, B.T. Ji, Q. Li, X.O. Shu, and Y.T. Gao, *The Shanghai Women's Health Study: rationale, study design, and baseline characteristics.* Am J Epidemiol, 2005. **162**(11): p. 1123-31.

2.	Gentleman, R.C., V.J. Carey, D.M. Bates, B. Bolstad, M. Dettling, S. Dudoit, B. Ellis, L. Gautier, Y. Ge, J. Gentry, K. Hornik, T. Hothorn, W. Huber, S. Iacus, R. Irizarry, F. Leisch, C. Li, M. Maechler, A.J. Rossini, G. Sawitzki, C. Smith, G. Smyth, L. Tierney, J.Y.H. Yang, and J. Zhang, *Bioconductor: open software development for computational biology and bioinformatics.* Genome Biology, 2004. **5**(10): p. R80.

**Supplemental Figure 1:** Exposure–response relationships between DNAm-IRS and lung cancer risk estimated using generalized additive models (binomial logit). Solid lines show odds ratios (ORs) relative to the median score (OR = 1), adjusted for age and BMI (held at median values). Dashed lines represent 95% confidence intervals. Curves are shown over approximately the 5th–95th percentile range of each score.

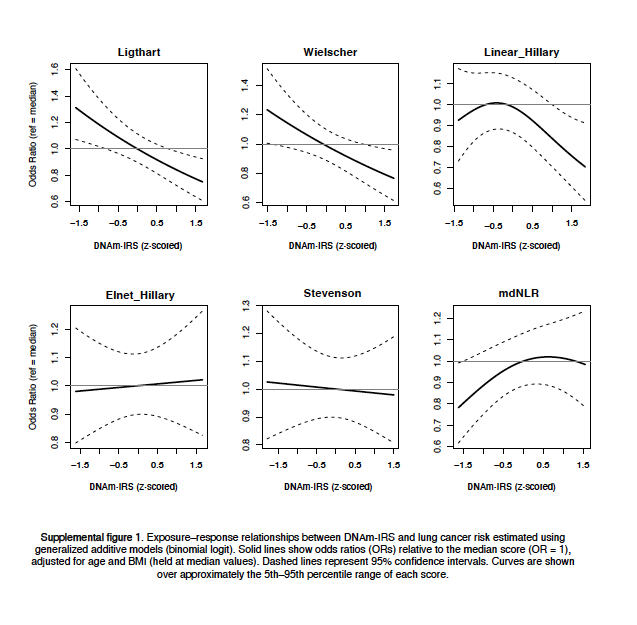

**Supplementary Figure 2. Weighted partial correlations of selected CRP-related DNAm scores with circulating protein modules and constituent markers.**
Weighted partial Spearman correlation coefficients were calculated between each DNAm-IRS and circulating protein module scores and constituent markers among participants with available immune-inflammatory marker data. Correlations were estimated under Model 1, adjusted for age and BMI, and Model 2, additionally adjusted for estimated leukocyte cell-type proportions. Rows show individual proteins and module scores, columns show DNAm-IRSs, and colored side bars denote protein module membership. Numbers within cells are weighted partial Spearman correlation coefficients, and color indicates the strength and direction of association. Asterisks indicate statistical significance: *P* < 0.05, **P** < 0.01, ***P*** < 0.001.

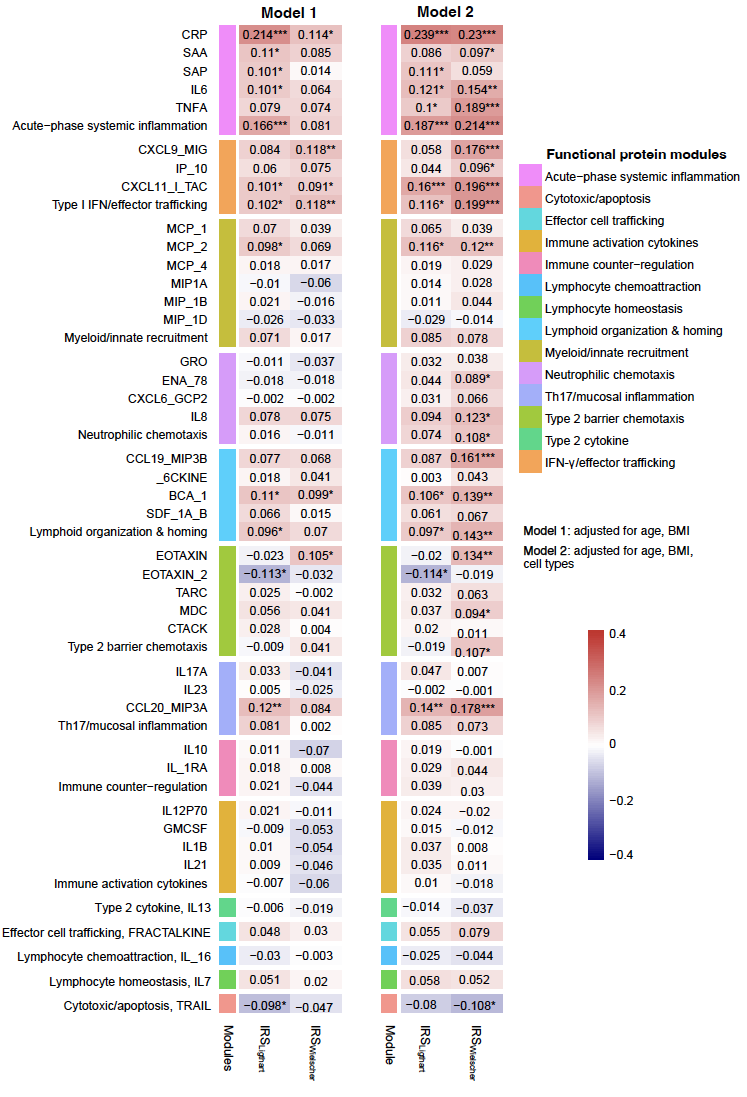
